# Vaccine effectiveness of a bivalent respiratory syncytial virus (RSV) pre-F vaccine against RSV-associated hospitalisation among adults aged 75-79 years in England

**DOI:** 10.1101/2025.06.13.25329583

**Authors:** Rebecca Symes, Heather J Whitaker, Shazaad Ahmad, David Arnold, Suryabrata Banerjee, Cariad M Evans, Robin Gore, Jennifer Hart, Katy Heaney, Onn Min Kon, Anne Melhuish, Munira Ortale Zogaib, Emanuela Pelosi, Najib Rahman, Gerrit Woltmann, Tricia McKeever, Maria Zambon, Conall H Watson, Wei Shen Lim, Jamie Lopez Bernal, the HARISS network collaborators

## Abstract

**Background:** A respiratory syncytial virus (RSV) vaccination programme for older adults using bivalent pre-F vaccine was introduced in England from September 1, 2024. While vaccine effectiveness has been reported against all-cause RSV-associated respiratory hospitalisations, there are limited data on vaccine effectiveness (VE) against different presentations of RSV-associated illness, such as exacerbation of chronic illness.

**Methods:** We conducted a test-negative design analysis using data from a national, hospital-based acute respiratory infection (ARI) sentinel surveillance (HARISS) system across 14 hospitals in England. From October 1, 2024 to March 31, 2025 vaccine-eligible adults aged 75-79 years hospitalised with ARI for ≥24 hours and tested with molecular diagnostic assays within 48 hours of admission were included. Cases were RSV positive, and controls negative for RSV, influenza and SARS-CoV-2. Vaccination status was obtained from the national registry.

**Findings:** Of 1006 adults hospitalised with ARI, 173 (17.2%) had RSV infection. VE was 82.3% (95%CI:70.6-90.0%) against hospitalisation for any RSV-associated ARI and 86.7% (75.4-93.6%) in those with severe disease including oxygen supplementation. VE was 88.6% (75.6-95.6%) among adults admitted due to lower respiratory tract infection (LRTI), including pneumonia, 77.4% (42.4-92.8%) among adults admitted due to exacerbation of chronic lung disease, and 78.8% (47.8-93.0%) among adults admitted with exacerbation of chronic heart disease, lung disease or frailty. In adults with immunosuppression, VE was 72.8% (39.5-89.3%).

**Interpretation:** This study provides evidence that the RSV pre-F vaccine is highly effective against RSV-associated hospitalisations, including exacerbations of chronic disease and in adults living with immunosuppression.

**Funding:** The UK Health Security Agency.

## Introduction

Respiratory syncytial virus (RSV) is a common seasonal respiratory virus that is an important cause of acute respiratory infection (ARI) in older adults in the UK, resulting in hospitalisation and mortality.^1-3^ Previous UK studies have estimated an annual hospital admission rate of 71 per 100,000 persons in adults aged 65-74 years increasing to 251 per 100,000 in adults aged 75 years and above.^3^ Those living with frailty and comorbidities such as chronic obstructive pulmonary disease (COPD) and chronic heart disease are at particularly high risk of severe clinical outcomes from RSV infection.^4-6^

From September 1, 2024 an RSV immunisation programme for older adults was introduced in England. The programme offers a single dose of bivalent RSV pre-F vaccine (Abrysvo, Pfizer, New York, USA) to all adults turning 75 years old alongside a one-off catch-up campaign for those aged 75 to 79 years old.^7^ The overall vaccine coverage in the catch-up cohort in England reached 60.3% as of March 31, 2025.^8^

Clinical trials report safety and efficacy against lower respiratory tract disease of the RSV pre-F vaccine but are limited by power to assess efficacy against severe outcomes such as hospitalisation and oxygen supplementation. Older adults with comorbidities including immunosuppression were also under-represented in these trials.^9-11^ Real world data from the US, where RSV vaccination was introduced in 2023, have demonstrated substantial vaccine effectiveness against RSV-associated hospitalisation.^12-16^ There is also early evidence of a population level impact of RSV vaccination in the UK.^17-18^ However, there are currently limited studies assessing the individual level effectiveness of RSV vaccine outside of the US.

Additionally, published studies do not report vaccine effectiveness against RSV-associated exacerbations of chronic illnesses such as lung disease or heart disease, common reasons for admission in older adults with RSV infection.^5-6^ Such presentations form a major component of the RSV burden in older adults^1^, therefore understanding the effectiveness against different types of hospitalisations is important when assessing the overall impact of RSV vaccination programmes. Furthermore, there is limited evidence on the effectiveness of RSV vaccination in different clinical risk groups, including immunosuppressed patients. In this study we seek to expand this evidence base using data from a national, hospital-based ARI sentinel surveillance (HARISS) system in England to report on end of season vaccine effectiveness from the first year of vaccine introduction in the UK. HARISS is an active surveillance system designed to collect detailed clinical data from hospital sites, enabling the assessment of vaccine effectiveness across a range of admission reasons.

This study aimed to estimate the effectiveness of RSV vaccination against hospitalisation due to RSV-associated ARI in England during the 2024/25 RSV season in vaccine-eligible older adults aged 75 to 79 years on September 1, 2024. We assessed vaccine effectiveness by severity of illness, by reason for admission, and by comorbidity status, including immunosuppression.

## Methods

### Study design

This case-control study, using a test negative design^19,20^, utilises data from HARISS, a national, hospital-based ARI sentinel surveillance (HARISS) system across 14 sentinel National Health Service (NHS) acute hospitals in England. HARISS is an active, enhanced surveillance system introduced in England in 2023/4 and expanded during 2024/5 from 7 to 14 hospital trust sites.^1^ Sites within the surveillance network include geographical representation across England (Supplementary figure 1). The study includes adults presenting to hospital from October 1, 2024 to March 31, 2025.

This surveillance was undertaken under Regulation 3 of The Health Service (Control of Patient Information) Regulations 2002 (UK legislation)^21^ which allows for collection of data for public health purposes without the requirement for individual informed patient consent. The UK Health Security Agency (UKHSA) Caldicott Advisory Panel and Research Ethics and Governance Group considered this study epidemiological surveillance and granted approval for the study to take place.

### Exposures

Individuals were classified as vaccinated if an RSV vaccination was administered at least 14 days prior to presentation to hospital, as part of the UK’s RSV immunisation programme.

Partially vaccinated individuals who received vaccine 0-13 days prior to presentation to hospital were excluded. Individuals were classified as unvaccinated if they had not received an RSV vaccination at the time of presentation to hospital. The UK’s RSV vaccination programme for older adult uses the bivalent Abrysvo pre-F vaccine, containing RSV-A and RSV-B derived antigen.

### Outcomes

The primary outcome was hospitalisation due to RSV-associated ARI. All HARISS sites were advised to test nasopharyngeal or combined nose and throat swabs for RSV, influenza (A and B) and SARS-CoV-2 in patients aged ≥65 years admitted to hospital for ≥24 hours where the reason for admission was symptomatic ARI. Symptomatic ARI included patients presenting with the following suspected diagnoses on admission: pneumonia, or pneumonitis; non-pneumonia lower respiratory tract infection (LRTI), or acute bronchitis; exacerbation of chronic lung disease e.g. COPD or asthma; exacerbation of chronic heart disease e.g. heart failure; exacerbation of frailty, or poor mobility e.g. fall; ARI with another reason for admission (clinical data collection questionnaire in supplementary file).

Cases were defined as adults admitted to hospital for at least 24 hours due to ARI, were positive for RSV on molecular diagnostic testing of samples taken within 48 hours of admission, and eligible for RSV vaccine on September 1, 2024 (aged 75-79 years). Cases were excluded from the main analysis if they had a coinfection with influenza or SARS-CoV-2, but included in a sensitivity analysis.

Controls were defined as adults admitted to hospital for at least 24 hours due to ARI, were negative for RSV, influenza and SARS-CoV-2 on molecular diagnostic testing of samples taken within 48 hours of admission, and eligible for RSV vaccine on September 1, 2024 (aged 75-79 years). For every RSV positive individual identified by the HARISS sites, a control was selected from the same epidemiological week. This ensured controls were selected throughout the RSV season, with more controls selected during periods of high RSV activity.

For individuals with two admissions for RSV, only the first admission for RSV was included. For individuals with multiple admissions for test-negative ARI, one admission was randomly selected for inclusion. If an individual was admitted for RSV and test-negative ARI, the RSV positive admission was included in the study. Individuals were excluded from the study if they were not identified in the Immunisation Information System (IIS) registry.

### Data sources

Clinical and laboratory data including reason for admission (multiple reasons selected if applicable), date of presentation to hospital, respiratory virus testing results and clinical outcomes during admission were captured from patient’s medical records using the Snap Survey platform (clinical data collection questionnaire in supplementary file). Medical records were reviewed by trained staff employed at each site. Clinical outcomes were captured at 30 days after sample date, or at discharge, whichever was sooner.

The IIS provided data on RSV vaccination status and date of administration, sex, ethnicity, Index of Multiple Deprivation (IMD), death date (where applicable) and influenza^22^ and COVID-19 clinical risk groups^23^ (to assess comorbidity status). The IIS is a national vaccine register containing vaccination and demographic information on the population of England registered with a General Practitioner. Comorbidities, including immunosuppression, are identified using the NHS CaaS (Cohorting as a Service), developed for call-recall purposes to identify at risk individuals requiring influenza or COVID-19 vaccine based on electronic health records.^24^ Immunosuppression includes individuals with immunosuppression due to disease or treatment as defined in the UK’s Immunisation against infectious disease Green Book.^22,23^ (Full list in supplementary file)

### Analysis

To assess the effectiveness of RSV vaccination against hospitalisation for RSV-associated ARI a test-negative case control analysis was conducted to compare the odds of vaccination among cases and controls using multivariable logistic regression. Vaccine effectiveness was calculated using 1-Odds Ratio, with a 95% confidence interval. The final multivariable logistic regression model was adjusted for days from start of the surveillance period to hospital presentation using splines, and presence of at least one comorbidity and/or immunosuppression. Other potential confounders examined for inclusion in the adjusted model were age, sex, ethnicity, index of multiple deprivation and region. These potential confounders did not change the vaccine effect by 1% or more and were therefore not included in the final adjusted model, as per protocol.

Vaccine effectiveness was estimated among adults with severe disease. Severe disease was denoted by the need during admission of: oxygen supplementation, high-flow nasal oxygen, non-invasive ventilation (NIV) or continuous positive airway pressure (CPAP), invasive ventilation or mechanical ventilation, and intensive care unit (ICU) admission; as well as mortality within 30 days of admission to hospital.

Vaccine effectiveness was estimated according to the following reasons for admission: LRTI including pneumonia; exacerbation of chronic lung disease (adults admitted with chronic lung disease exacerbation but excluding those with LRTI); and exacerbation of chronic lung disease, heart disease and/or frailty (adults admitted with any of these diagnoses, excluding those with LRTI).

Vaccine effectiveness was also stratified by comorbidity status: without immunosuppression; without immunosuppression, liver disease and renal disease; with immunosuppression; with chronic respiratory disease; and with chronic heart and vascular disease. A sensitivity analysis was conducted to explore the inclusion of cases that tested positive for influenza (A or B) and SARS-CoV-2 coinfection. To reduce the inclusion of underpowered results, vaccine effectiveness estimates were excluded if any expected cell count for vaccinated or unvaccinated cases or controls was less than 10.

Demographic characteristics and comorbidities were described for cases and controls. Mann-Whitney U tests, Pearson’s chi-squared tests and Fisher’s exact tests, where applicable, were used to compare demographic and comorbidities characteristics between groups. P-values of <0.05 were regarded as significant.

Missing data were minimised by using compulsory fields in the Snap Survey data collection tool. Individuals were included in the study only if identified in the IIS dataset. Missing data e.g. for ethnicity and IMD has been reported. Analyses were completed using R version 4.3.2.^25^

## Results

Between October 1, 2024 and March 31, 2025, 1006 hospital admissions from 14 HARISS network sites were included in this study (Figure 1); 173 (17.2%) cases and 833 (82.8%) controls. Peak RSV positivity admissions occurred week beginning November 18, 2024, with the second highest activity week beginning December 16, 2024 (Figure 2).

**Figure 1:**
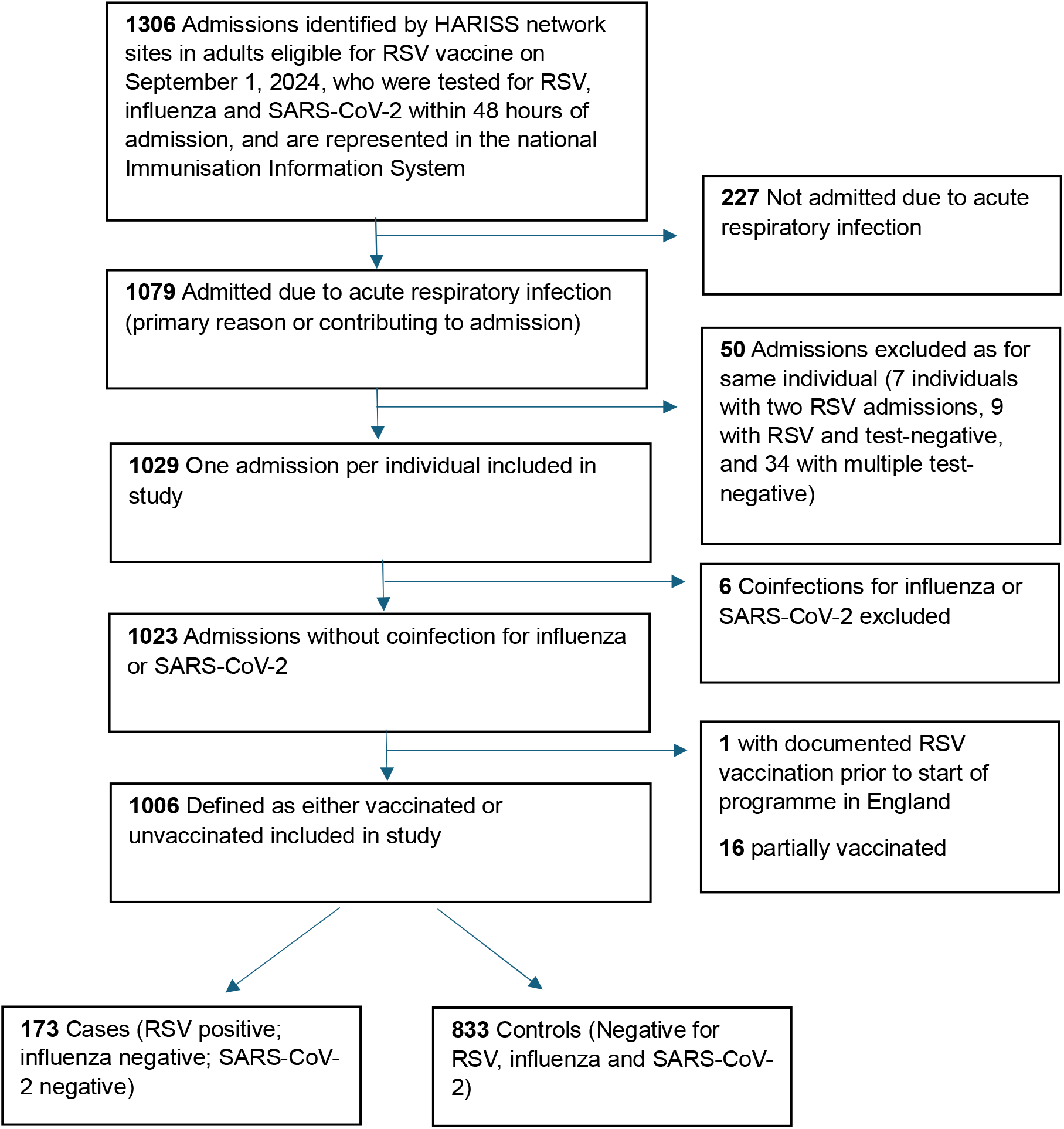
Study flow diagram

**Figure 2:**
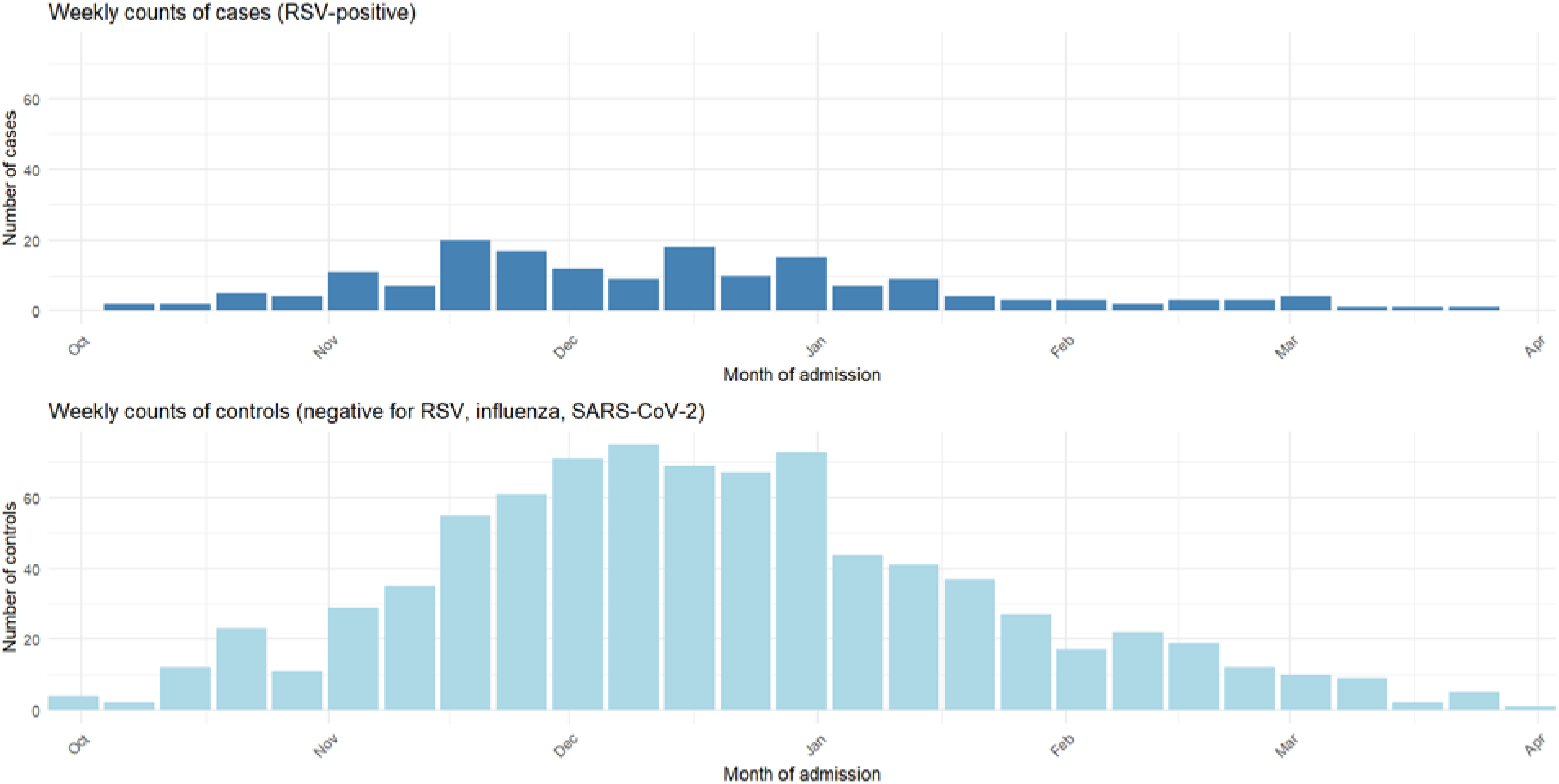
Epidemiological curve of cases and controls by week of presentation to hospital, between October 1, 2024 and March 31, 2025

The median age for both case and control groups was 77 years (IQR 76-78). There were similar proportions of females among cases and controls (53.8% vs 52.0%) and the majority of adults were of White ethnicity (89.6% in cases and 88.8% in controls). Adults were from the most deprived IMD quintile in 23.7% of cases and 26.3% of controls. The majority of adults had at least one medical comorbidity (cases 90.8%; controls 92.4%).

Immunosuppression was present in 27.2% of cases and 26.9% of controls. High proportions of cases and controls had chronic respiratory disease (53.8% vs 61.0%) and chronic heart and vascular disease (63.6% vs 60.7%) (Table 1).

**Table 1:** Demographic characteristics and comorbidities for HARISS cohort by cases and controls.

| **N (%), unless otherwise stated** | **Cases** | **Controls** | **p-value** |
| --- | --- | --- | --- |
| Demographics |  |  |  |
| Total | 173 (17.2) | 833 (82.8) |  |
| Age (median (IQR)) | 77 (76-78) | 77 (76-78) | 0.13 |
| Sex (female) | 93 (53.8) | 433 (52.0) | 0.67 |
| Ethnicity |  |  | 0.97 |
| White | 155 (89.6) | 740 (88.8) |  |
| Mixed or multiple ethnic groups | 0 (0) | 3 (0.4) |  |
| Asian or Asian British | 6 (3.5) | 36 (4.3) |  |
| Black, Black British, Caribbean or African | 1 (0.6) | 10 (1.2) |  |
| Other ethnic group | 2 (1.2) | 12 (1.4) |  |
| Missing ethnicity | 9 (5.2) | 32 (3.8) |  |
| Index of Multiple Deprivation (IMD) quintile |  |  | 0.35 |
| 1 (Most deprived) | 41 (23.7) | 219 (26.3) |  |
| 2 | 22 (12.7) | 141 (16.9) |  |
| 3 | 36 (20.8) | 131 (15.7) |  |
| 4 | 36 (20.8) | 157 (18.8) |  |
| 5 (Least deprived) | 37 (21.4) | 181 (21.7) |  |
| Missing IMD | 1 (0.6) | 4 (0.5) |  |
| Comorbidities |  |  |  |
| At least one comorbidity | 157 (90.8) | 770 (92.4) | 0.45 |
| Chronic respiratory disease | 93 (53.8) | 508 (61.0) | 0.08 |
| Chronic neurological disease | 48 (27.7) | 227 (27.3) | 0.89 |
| Chronic liver disease | 4 (2.3) | 25 (3.0) | 0.80 |
| Chronic kidney disease | 46 (26.6) | 213 (25.6) | 0.78 |
| Chronic heart disease and vascular disease | 110 (63.6) | 506 (60.7) | 0.49 |
| Immunosuppression | 47 (27.2) | 224 (26.9) | 0.94 |
| Diabetes and other endocrine disorders | 51 (29.5) | 242 (29.1) | 0.91 |
| Morbid obesity | 6 (3.5) | 42 (5.0) | 0.38 |
| Severe mental illness | 8 (4.6) | 25 (3.0) | 0.28 |
| RSV vaccination status |  |  |  |
| Vaccinated | 16 (9.2) | 308 (37.0) | <0.0001 |

Note: p-values calculated using Mann-Whitney U tests, Pearson's chi-squared tests and Fisher's exact tests where applicable

Among all included admissions, 324 (32.2%) adults had received an RSV vaccination 14 days or more prior to presentation to hospital with an ARI; comprising 16 (9.2%) cases and 308 (37.0%) controls (Table 1). The main difference in demographic characteristics between vaccinated and unvaccinated groups was sex; 47.2% female in vaccinated and 54.7% in unvaccinated (p=0.027). The majority of patients were of White ethnicity (vaccinated 90.1%; unvaccinated 88.4%), and 23.5% of those vaccinated and 27.0% of those unvaccinated were in the most deprived IMD quintile (Table 2).

**Table 2:** Demographic characteristics and comorbidities for the HARISS cohort by vaccination status.

| **N (%), unless otherwise stated** | **Vaccinated** | **Unvaccinated** | **p-value** |
| --- | --- | --- | --- |
| Demographics |  |  |  |
| Total | 324 (32.2) | 682 (67.8) |  |
| Age (median (IQR)) | 77 (76-78) | 77 (76-78) | 0.13 |
| Sex (female) | 153 (47.2) | 373 (54.7) | 0.027 |
| Ethnicity |  |  | 0.88 |
| White | 292 (90.1) | 603 (88.4) |  |
| Mixed or multiple ethnic groups | 1 (0.3) | 2 (0.3) |  |
| Asian or Asian British | 12 (3.7) | 30 (4.4) |  |
| Black, Black British, Caribbean or African | 2 (0.6) | 9 (1.3) |  |
| Other ethnic group | 4 (1.2) | 10 (1.5) |  |
| Missing ethnicity | 13 (4.0) | 28 (4.1) |  |
| Index of Multiple Deprivation (IMD) quintile |  |  | 0.11 |
| 1 (Most deprived) | 76 (23.5) | 184 (27.0) |  |
| 2 | 49 (15.1) | 114 (16.7) |  |
| 3 | 46 (14.2) | 121 (17.7) |  |
| 4 | 69 (21.3) | 124 (18.2) |  |
| 5 (Least deprived) | 83 (25.6) | 135 (19.8) |  |
| Missing IMD | 1 (0.3) | 4 (0.6) |  |
| Comorbidities |  |  |  |
| At least one comorbidity | 302 (93.2) | 625 (91.6) | 0.39 |
| Chronic respiratory disease | 204 (63.0) | 397 (58.2) | 0.15 |
| Chronic neurological disease | 80 (24.7) | 195 (28.6) | 0.19 |
| Chronic liver disease | 8 (2.5) | 21 (3.1) | 0.59 |
| Chronic kidney disease | 84 (25.9) | 175 (25.7) | 0.93 |
| Chronic heart disease and vascular disease | 202 (62.3) | 414 (60.7) | 0.62 |
| Immunosuppression | 97 (29.9) | 174 (25.5) | 0.14 |
| Diabetes and other endocrine disorders | 95 (29.3) | 198 (29) | 0.92 |
| Morbid obesity | 15 (4.6) | 33 (4.8) | 0.38 |
| Severe mental illness | 8 (2.5) | 25 (3.7) | 0.32 |

Note: p-values calculated using Mann-Whitney U tests, Pearson's chi-squared tests and Fisher's exact tests where applicable

Among older adults aged 75-79 years on 1 September 2024, the effectiveness of RSV vaccination against hospitalisation for any RSV-associated ARI was 82.3% (95% CI 70.6-90.0%). Vaccine effectiveness against any RSV-associated ARI amongst those with severe disease was 86.7% (95% CI 75.4-93.6%). Among adults with severe disease, 78.8% received oxygen supplementation alone.

Vaccine effectiveness was 88.6% (95% CI 75.6-95.6%) among adults admitted due to a lower respiratory tract infection (LRTI), including pneumonia; 77.4% (95% CI 42.4-92.8%) among adults admitted due to an exacerbation of chronic lung disease such as COPD; and 78.8% (95% CI 47.8-93.0%) among those admitted for an exacerbation of any chronic illness including chronic lung disease, chronic heart disease and/or frailty (Figure 3).

**Figure 3:**
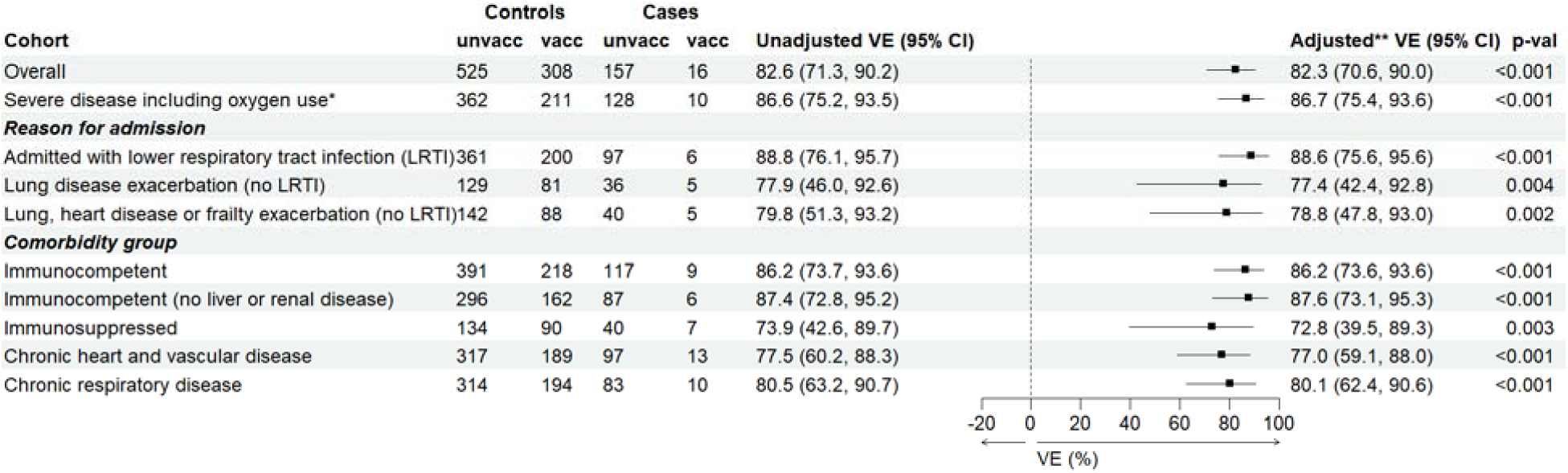
Estimated vaccine effectiveness (VE) against hospitalisation for respiratory syncytial virus-associated acute respiratory infection, in the HARISS network cohort, 2024/5 Notes: *Severe disease includes adults indicated as requiring during admission: oxygen supplementation, high-flow nasal oxygen, non-invasive ventilation (NIV) or continuous positive airway pressure (CPAP), invasive ventilation or mechanical ventilation, and intensive care unit (ICU) admission, and adults that died within 30 days of admission to hospital. **Vaccine effectiveness adjusted for days from start of the surveillance period to hospital presentation using splines, and presence of at least one comorbidity and/or immunosuppression

When assessed according to underlying comorbid illnesses, vaccine effectiveness was 86.2% (95% CI 73.6-93.6%) in immunocompetent individuals, 87.6% (95% CI 73.1-95.3%) in immunocompetent individuals without hepatic or renal disease, and 72.8% (95% CI 39.5-89.3%) in individuals with immunosuppression. Vaccine effectiveness was 77.0% (95% CI 59.1-88.0%) in those with chronic heart and vascular disease and 80.1% (95% CI 62.4-90.6%) in those with chronic respiratory disease (Figure 3).

A sensitivity analysis that included RSV positive cases with either influenza or SARS-CoV-2 coinfection (n=6) demonstrated no meaningful changes in vaccine effectiveness against hospitalisation due to RSV-associated ARI (supplementary figure 2).

## Discussion

This study utilising data from an active, national, hospital-based enhanced surveillance network in England (HARISS) demonstrates real-world effectiveness of the RSV pre-F vaccine (Abrysvo) against hospitalisation due to RSV-associated ARI in older adults. We found vaccine effectiveness against hospitalisation for any RSV-associated ARI in adults aged 75-79 years to be 82% in the first season since immunisation programme introduction in the UK. The vaccine demonstrated effectiveness of 77-89% across different presentations of RSV-associated infection, including those admitted due to exacerbations of chronic disease. Vaccine protection was also maintained across different comorbidity groups including those with immunosuppression.

To our knowledge, our findings are among the first to demonstrate RSV vaccine effectiveness against hospitalisation in older adults in Europe, with the potential to impact policy in the UK and other regions with similar healthcare structures. Additionally, these findings provide new evidence on RSV vaccine effectiveness in adults admitted due to RSV-associated chronic illness exacerbations such as exacerbations of COPD. These additional data are important to direct vaccine policy and support cost-effectiveness evaluations, particularly for programmes that might be focused on individuals living with major co-morbid conditions.

The RENOIR clinical trial for the RSV pre-F vaccine reported vaccine efficacy of 85.7% (96.66% CI 32.0-98.7) against RSV with at least three lower respiratory symptoms or signs, during the first season.^9^ Although there are differences in the endpoint studied and population characteristics, such as higher levels of chronic medical conditions in our cohort, our findings are consistent with the trial data, with both studies demonstrating significant vaccine protection.

Furthermore, real world studies of RSV vaccine effectiveness conducted in the US have reported effectiveness against hospitalisation in older adults of 80.3% (95% CI 65.8– 90.1%)^13^, 75.5% (95% CI 73.1-77.6%)^16^, 75% (95% CI 50-87%)^15^, 91% (95% CI 59-98%) where emergency department visits were also included^12^, and 80% (95% CI 71–85%) in immunocompetent and 73% (95% CI 48–85%) in immunosuppressed older adults^14^. Vaccine effectiveness specifically for the RSV pre-F vaccine has been reported at 73% (95% CI 52– 85%) against RSV-associated hospitalisation^14^, 77.4% (95% CI 70.4-82.4%) against RSV infections^13^, and 91% (95% CI, 59%-98%) against RSV-related lower respiratory tract disease hospitalisation or emergency department visits.^12^ Our overall estimate of vaccine effectiveness is consistent with these previous studies, with overlapping confidence intervals. Although our study focuses on individuals aged 75-79 years only, in other studies, vaccine effectiveness across different age groups (60-74 years and ≥75 years) has been reported to be similar.^14,15^

When focussing on more severe disease including hospitalisations requiring oxygen supplementation, the point estimate for vaccine effectiveness was higher at 86.7% (95% CI 75.4-93.6%), though confidence intervals overlapped with the overall vaccine effectiveness estimates. In one other study assessing RSV vaccine effectiveness amongst adults with severe disease, Tartof *et al*.^12^ found a vaccine effectiveness of 89% (95% CI 13-99%) in severe RSV-related lower respiratory tract disease, compared to 91% (95% CI 59-98%) in the whole cohort, although with wide and overlapping confidence intervals. In contrast to our study, oxygen use was the only disease severity outcome used. For other respiratory virus vaccines, such as the COVID-19 vaccine, higher vaccine effectiveness against more severe disease has been observed.^26^

Exacerbations of chronic disease, especially chronic heart or lung disease, accounts for a large proportion of hospitalisations due to RSV infection.^5,6^ Tsueng *et al*. report 80% of older adults with COPD/chronic bronchitis/emphysema experienced an exacerbation of these chronic lung diseases during hospitalisation for RSV infection^27^, emphasising a key opportunity for prevention of severe illness through vaccination. This study provides evidence of vaccine effectiveness both in older adults admitted due to LRTI including pneumonia, and in those admitted due to RSV-associated exacerbations of chronic illness. Of note, while vaccine effectiveness remained high, point estimates were lower in patients presenting with exacerbations of chronic lung disease (77.4%, 95% CI 42.4-92.8), compared to those presenting with LRTI (88.6%, 95% CI 75.6-95.6), though confidence intervals overlapped. It will be important for future studies to determine if this potential difference in vaccine effectiveness is significant. The clinical impacts and outcomes of RSV-associated LRTI (including pneumonia) may differ compared to RSV-associated exacerbations of chronic illnesses, and differences in vaccine effectiveness according to these different presentations would influence future modelling and cost-effectiveness assessments.

Older adults with immunosuppression comprised a relatively large proportion of the study cohort (26.9%) underlining the risks from RSV infection faced by these individuals. We observed a vaccine effectiveness against hospitalisation of 72.8% amongst immunosuppressed individuals. This is comparable to results reported by Payne *et al*. (who observed vaccine effectiveness of 73%) in their study of US adults where the proportion of immunocompromised individuals in their cohort was similar at 23%.^14^

The substantial vaccine protection reported in this study underscores the importance of RSV vaccination in reducing severe outcomes experienced by at risk populations including older adults and those with chronic medical conditions.^28,29^

Strengths of this study include the use of a national, sentinel surveillance system for ARI with strong geographical representation across England, increasingly generalisability. HARISS is an active, enhanced surveillance system collecting specific data to meet this study’s aims. This surveillance system enables the collection of data on specific reasons for admission and outcomes amongst those who meet a case definition, and therefore does not rely on routine data sources. The use of the HARISS network, where hospitals are recruited due to consistent routine molecular testing for RSV, influenza and SARS-CoV-2 in older adults presenting with ARI, reduces bias caused by selective testing. The direct review of patient notes by trained staff resulted in detailed clinical information being obtained. Furthermore, the inclusion of only admissions with respiratory virus swabbing within 48 hours of presentation to hospital resulted in the inclusion of cases who had been admitted due to RSV-associated ARI rather than nosocomial infection. The use of a national vaccination registry provided reliable reports of vaccination status.

There are several limitations of this study. The size of the study cohort did not allow assessment of vaccine effectiveness against more severe outcomes alone (without oxygen supplementation) such as intensive care admission and death at 30 days. As the controls selected for this study were negative for RSV, influenza and SARS-CoV-2 we were unable to conduct a sensitivity analysis using controls negative for RSV but positive for influenza and/or SARS-CoV-2. Testing was conducted as part of routine clinical care and not for research purposes and therefore there may be some variation in testing procedures between sites. Not all respiratory samples were tested using an extended respiratory viruses panel e.g. for rhinovirus, we are unable to report this positivity information for the control group. Additionally, we were unable to report vaccine effectiveness by vaccine type as only one vaccine product has been used in England to date. Although we assessed relevant confounders and adjusted for these where appropriate in the analysis, there is the possibility of residual confounding, for example severity of chronic illness or other comorbidities such as musculoskeletal conditions, that is not currently captured in the comorbidity data. Lastly, we do not report subtyping data as this information was not available for all samples. Assuming the distribution of subtypes across the hospital population is similar to the wider community, both RSV-A and RSV-B subtypes were identified in similar proportions in General Practice all age data over winter 2024/25 in England.^30^ Further monitoring of vaccine effectiveness and genomic surveillance is warranted over different RSV seasons.

## Conclusions

This analysis is among the first to present evidence of real-world effectiveness of the RSV pre-F vaccine against RSV-associated hospitalisations in older adults in Europe, including in adults admitted due to exacerbations of chronic illness. The high vaccine effectiveness observed across different RSV-related presentations and in individuals with a range of underlying comorbid illnesses supports the promotion of high vaccine coverage to reduce RSV-associated illness in adults.

## Data Availability

Original data are confidential and no additional data available.

## HARISS network collaborators (co-authors)

Matthew Donati^a^, Elisabeth North^b^, Hongyi Zhang^c^, Chloe Myers^c^, Bethan Phillips^d^, Ajit Lalvani^e^, Paul Randell^f^, Joan Nanan^g^, Fiona McGill^h^, Christopher W Holmes^i,j^, Claire L. McMurray^i,k^, Samantha Brightmore^l^, Leanne Small^l^, Tim William Felton^m,n,o^, Nicholas Machin^p^, Brendan Payne^q^, Nikhil Premchand^r^, John Steer^r^, Louise Berry^s^, Louise Lansbury^t,u^, Tine Panduro^v,w^, Melissa Dobson^v,w^, Monique Andersson^x^, Marc Lipman^y^, Lise Ridge^z^, Thomas Swaine^aa^, Adam Hinchcliffe^aa^, Charlotte Si Yuan Lim^aa^, Aimee Serisier^aa^, Tamsin McKinnon^aa^, Simon Tazzyman^bb^, Thushan de Silva^cc^, Tristan W Clark^dd^, Christopher Rawlinson^ee^, Alec Cobbold^ee^

a. UK Health Security Agency, South West Regional Laboratory and Severn Infection Sciences, Bristol, UK

b. Academic Respiratory Unit, North Bristol NHS Trust, Bristol, UK

c. UKHSA Clinical Microbiology and Public Health Laboratory, Cambridge University Hospitals NHS Foundation Trust, Addenbrooke’s Hospital, Cambridge, UK

d. Frimley Health NHS Foundation Trust, Frimley, UK

e. NIHR Health Protection Research Unit in Respiratory Infections, National Heart and Lung Institute, Imperial College London, London, UK

f. Imperial College Healthcare NHS Trust, London, UK

g. National Heart and Lung Institute, Imperial College London and Imperial College Healthcare NHS Trust, London, UK

h. Leeds Teaching Hospitals NHS Trust, Leeds, UK

i. Department of Clinical Microbiology, University Hospitals of Leicester, UK

j. Department of Respiratory Sciences, University of Leicester, Leicester, UK

k. Department of Genetics, Genomics and Cancer Sciences, University of Leicester, Leicester, UK

l. University Hospitals of Leicester NHS Trust, Leicester, UK

m. The University of Manchester, Manchester, UK

n. NIHR Manchester Biomedical Research Centre, Manchester, UK

o. Manchester University NHS Foundation Trust, Manchester, UK

p. Virology Department, Manchester Medical Microbiology Partnership, Manchester University NHS Foundation Trust, UK

q. The Newcastle-upon-Tyne Hospitals NHS Foundation Trust, Newcastle, UK

r. Northumbria Healthcare NHS Foundation Trust, Newcastle, UK

s. Department of Microbiology, Nottingham University Hospitals NHS Trust, Nottingham, UK

t. Faculty of Medicine and Health Sciences, University of Nottingham, Nottingham, UK

u. National Institute for Health Research (NIHR) Nottingham Biomedical Research Centre, UK

v. Oxford Respiratory Trials Unit, University of Oxford, Oxford, UK

w. Nuffield Department of Clinical Medicine, University of Oxford, Oxford, UK

x. Nuffield Department of Clinical Laboratory Sciences, Radcliffe Department of Medicine, University of Oxford, Oxford, UK

y. UCL Respiratory, Division of Medicine UCL & Respiratory Medicine, Royal Free Hospital, Royal Free London NHS Foundation Trust, London

z. Respiratory Medicine, Royal Free Hospital, Royal Free London NHS Foundation Trust, London, UK

aa. Department of Virology, Royal Free Hospital, Royal Free London NHS Foundation Trust, London, UK

bb. South Yorkshire and Bassetlaw Pathology Network, Sheffield Teaching Hospitals NHS Trust, Sheffield, UK

cc. Division of Clinical Medicine, School of Medicine and Population Health, The University of Sheffield, Sheffield, UK

dd. School of Clinical and Experimental Sciences, University of Southampton and Department of Infection, University Hospitals Southampton NHS Foundation Trust, UK

ee. Immunisation and Vaccine-Preventable Diseases Division, UK Health Security Agency, London, UK

## Acknowledgements

*We would like to acknowledge the following colleagues for their contributions:*

*UK Health Security Agency, South West Regional Laboratory and Severn Infection Sciences, Bristol, UK: Paul North, Rich Hopes*

*Cambridge University Hospitals NHS Foundation Trust, UK: Chara Alexiou*

*UKHSA Clinical Microbiology and Public Health Laboratory, Cambridge University Hospitals NHS Foundation Trust Addenbrooke’s Hospital, UK: Pedro Costa*

*Frimley Health NHS Foundation Trust, UK: Helen A Powell, Reanna Allen Leeds Teaching Hospitals NHS Trust, UK: Tadas Mazeika, Richard Winetrobe*

*Department of Clinical Microbiology, University Hospitals of Leicester, Leicester Royal Infirmary, UK: Mina Odedra*

*Manchester University NHS Foundation Trust, Manchester, UK: Maria Swizel Furtado e Aguiar, Manju Juby, Oluwafunmilayo Adungba, Luke Ward, Georgios Chalikias*

*The Newcastle-upon-Tyne Hospitals NHS Foundation Trust, UK: Gary Eltringham Northumbria Healthcare NHS Foundation Trust, UK: Ruth Henein, Aimee Joyce, Michelle Forster, Fatima Babatunde*

*Nottingham University Hospitals NHS Trust, Nottingham, UK: Lesley Bendall, Depu Parakkalpapu, Michelle Lister, Benjamin Sloan*

*Oxford Respiratory Trials Unit, University of Oxford, Oxford, UK: Ravirekha Raguchandra, Jasmin Sadler-Ladell*

*Department of Virology, Royal Free Hospital, Royal Free London NHS Foundation Trust, UK: Douglas Fink, Tanzina Haque. Health Services Laboratories, London, UK: Melanie Turner Clinical pathology testing service for this work at Royal Free London was delivered by Health Services Laboratories a partnership between The Doctors Laboratory (TDL), the Royal Free London NHS Foundation Trust and University College London Hospitals NHS Foundation Trust*.

*Clinical Trials Assistant Team, Sheffield Teaching Hospitals NHS Trust, UK: Hannah Dunn, Jessica Gregory, Christopher Norman, Cameron Obie, Abigail Olojede, Jennifer Rufus, Franciszek Skalbania, Fred Smith, Nicole Whitfield, Chloe Mathewmann, Hannah Maughan, Georgie MacLoughlin, Nicola Tinker*

*Department of Virology, Sheffield Teaching Hospitals NHS Trust, UK: Alex Yates*

*University Hospital Southampton, NHS Foundation Trust, Southampton, UK: Sophie Willis, Chibuzor Amechi, Nicola White*

*UKHSA, London: Thomas Williams*

***We would like to acknowledge the NIHR Respiratory Biomedical Research Centres and NIHR Respiratory Translational Research Collaboration***.

