## Supplementary file for "Vaccine effectiveness of a bivalent respiratory syncytial virus (RSV) pre-F vaccine against RSV-associated hospitalisation among adults aged 75-79 years in England"

### Contents

**Supplementary Figure 1: Map of England with location of HARISS sites…………………page 2**

**Supplementary Figure 2: Vaccine effectiveness sensitivity analysis including RSV-positive cases with coinfection for influenza or SARS-CoV-2………………page 2**

**Supplementary material: Clinical risk group (Immunosuppression)………………………..page 3**

**Supplementary material: Data collection questionnaire……………………………………...page 4**

**Supplementary Figure 1: Map of England with location of HARISS sites**

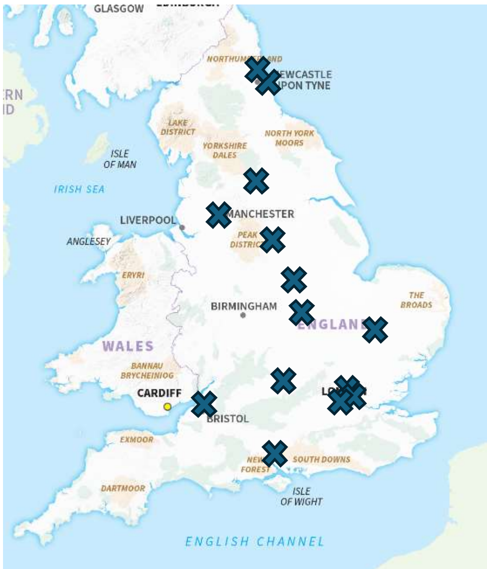

Acknowledgement: Ordnance Survey January 2025. Accessed from: GB Overview Maps | Data Products | OS. Adapted to show HARISS sites location.

**Supplementary Figure 2: Vaccine effectiveness sensitivity analysis including RSV-positive cases with coinfection for influenza or SARS-CoV-2**

n=1012 (6 additional RSV-positive cases with coinfection for influenza or SARS-CoV-2)

| | Controls | | Cases | | | | |
|---|---|---|---|---|---|---|---|
| Cohort | unvacc | vacc | unvacc | vacc | Unadjusted VE (95% CI) | Adjusted** VE (95% CI) | p-val |
| Overall | 525 | 308 | 162 | 17 | 82.1 (70.8, 89.7) | 81.8 (70.2, 89.6) | <0.001 |
| Severe disease including oxygen use* | 362 | 211 | 133 | 11 | 85.8 (74.3, 92.9) | 86.0 (74.6, 93.0) | <0.001 |
| ***Reason for admission*** | | | | | | | |
| Admitted with lower respiratory tract infection (LRTI) | 361 | 200 | 102 | 6 | 89.4 (77.3, 95.9) | 89.3 (77.0, 95.9) | <0.001 |
| Lung disease exacerbation (no LRTI) | 129 | 81 | 36 | 5 | 77.9 (46.0, 92.6) | 77.4 (42.4, 92.8) | 0.004 |
| Lung, heart disease or frailty exacerbation (no LRTI) | 142 | 88 | 40 | 5 | 79.8 (51.3, 93.2) | 78.8 (47.8, 93.0) | 0.002 |
| ***Comorbidity group*** | | | | | | | |
| Immunocompetent | 391 | 218 | 118 | 10 | 84.8 (71.8, 92.7) | 84.8 (71.6, 92.7) | <0.001 |
| Immunocompetent (no liver or renal disease) | 296 | 162 | 87 | 7 | 85.3 (69.6, 93.9) | 85.5 (69.8, 94.0) | <0.001 |
| Immunosuppressed | 134 | 90 | 44 | 7 | 76.3 (48.2, 90.6) | 75.1 (45.1, 90.2) | 0.001 |
| Chronic heart and vascular disease | 317 | 189 | 99 | 14 | 76.3 (58.6, 87.3) | 75.7 (57.6, 87.1) | <0.001 |
| Chronic respiratory disease | 314 | 194 | 87 | 10 | 81.4 (65.0, 91.1) | 81.1 (64.3, 91.0) | <0.001 |

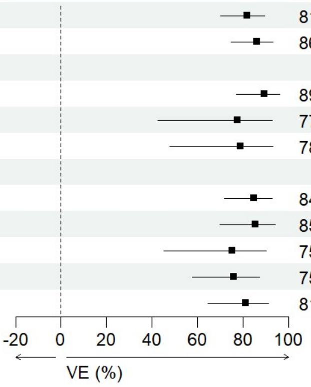

Notes:

*Severe disease includes adults indicated as requiring during admission: oxygen supplementation, high-flow nasal oxygen, non-invasive ventilation (NIV) or continuous positive airway pressure (CPAP), invasive ventilation or mechanical ventilation, and intensive care unit (ICU) admission, and adults that died within 30 days of admission to hospital.

**Vaccine effectiveness adjusted for days from start of the surveillance period to hospital presentation using splines, and presence of at least one comorbidity and/or immunosuppression

**Supplementary material: Clinical risk group (Immunosuppression)** as per the UK's Immunisation against infectious diseases Green Book for Influenza and COVID-19

Immunosuppression due to disease or treatment, including:

- Patients undergoing chemotherapy leading to immunosuppression.
- Patients undergoing radical radiotherapy.
- Solid organ transplant recipients, bone marrow or stem cell transplant recipients.
- HIV infection at all stages.
- Multiple myeloma or genetic disorders affecting the immune system (e.g. IRAK-4, NEMO, complement disorder, SCID).
- Individuals who are receiving immunosuppressive or immunomodulating biological therapy including, but not limited to, anti-TNF, alemtuzumab, ofatumumab, rituximab, patients receiving protein kinase inhibitors or PARP inhibitors, and individuals treated with steroid sparing agents such as cyclophosphamide and mycophenolate mofetil.
- Individuals treated with or likely to be treated with systemic steroids for more than a month at a dose equivalent to prednisolone at 20mg or more per day for adults.
- Anyone with a history of haematological malignancy, including leukaemia, lymphoma, and myeloma.
- Those who require long term immunosuppressive treatment for conditions including, but not limited to, systemic lupus erythematosus, rheumatoid arthritis, inflammatory bowel disease, scleroderma and psoriasis.

**Supplementary material: Data collection questionnaire**

| | |
|---|---|
| | **Questionnaire (ARI surveillance)** |
| | NHS number, name, date of birth |
| | Date of respiratory sample (*enter date*). Respiratory sample results (tick): *RSV positive, Influenza A positive, Influenza B positive, SARS-CoV-2 positive, positive for another respiratory pathogen, negative for all respiratory pathogens tested* |
| | Was this patient admitted to hospital for ≥24 hours? Yes – completes rest of questionnaire. No – does not complete rest of questionnaire. |
| | **Date of presentation to hospital** |
| **1** | Date of presentation to hospital (please enter the date of presentation to hospital for this episode of illness) |
| | *Calendar box to enter date* |
| | **Symptoms** |
| **2a** | Did the patient report or present with respiratory symptoms or fever on admission? |
| | *Yes/No/Unknown* |
| **2b/c** | Date of onset of first respiratory symptoms or fever (or symptoms that led to admission if no respiratory symptoms or fever) |
| | *Calendar box to enter date* |
| **2b** | Did the patient report or present with any of the following signs and symptoms on admission? (Please tick all that apply) |
| | *Documented fever (≥37.8 C) or history of fever* |
| | *Hypothermia (<35.5 C)* |
| | *New or increased cough* |
| | *New or increased sputum volume or discolouration* |
| | *New or increased shortness of breath* |
| | *New or increased wheezing* |
| | *Respiratory rate ≥25/min* |
| | *The patient did not present or report any of the above signs or symptoms on admission* |
| | *Unknown* |
| | **Reason for admission** |
| **3a** | Was the patient admitted because of a symptomatic acute respiratory infection? |
| | *Yes, symptomatic acute respiratory infection primary reason for admission (e.g. pneumonia)* |
| | *Yes, symptomatic acute respiratory infection contributing to admission (eg. RSV infection exacerbating heart failure)* |
| | *No, admission unrelated to symptomatic acute respiratory infection (eg. cellulitis)* |
| | If no, please give reason for admission |
| **3b** | The patient was admitted because of… (Please tick all that apply) |
| | *Pneumonia or pneumonitis* |
| | *Non-pneumonia lower respiratory infection or acute bronchitis* |
| | *Exacerbation of chronic lung disease (e.g. COPD)* |
| | *Exacerbation of chronic heart disease (e.g. heart failure/angina)* |
| | *Exacerbation of frailty or poor mobility* |
| | *Symptomatic acute respiratory infection with another reason for admission not specified above (eg. cough syncope) – please specify reason for admission (box provided)* |

| | |
|---|---|
| | **Antiviral use** |
| **4a** | Did the patient receive antivirals (for this illness episode) prior to admission? |
| **4b** | If yes, please specify which antiviral |
| | *Tick box for: Oseltamivir, Zanamivir, Paxlovid, Sotrovimab, Remdesivir, Other (please specify), Unknown* |
| | **Severity**** |
| **5a** | During this hospital admission, did the patient require any of the following: *(Please tick all that apply):* |
| | *Oxygen via cannulae or mask* |
| | *High-flow nasal oxygen (HFNO)* |
| | *Non-invasive ventilation (NIV) or continuous positive airway pressure (CPAP)* |
| | *Invasive ventilation or mechanical ventilation* |
| | *Intensive Care Unit (ICU) admission* |
| | *Extracorporeal membrane oxygenation (ECMO)* |
| | *None of the above were required during this hospital admission* |
| | *Unknown* |
| **5b** | Did the patient die during this admission? (If length of stay >30 days please record death outcomes up to 30 days since admission) |
| | *Yes/ No/ Unknown* |
| **5c** | Did the patient die as a result of (primary or contributory cause of death) an acute respiratory infection, or its complications? |
| | *Yes* |
| | *No* |
| | *Unknown* |
| **5d** | If the patient has died, please provide the cause of death as recorded on the death certificate or in the patient notes: |
| | *Cause of death 1a:* |
| | *Cause of death 1b:* |
| | *Unknown* |
| | **Further information** |
| | Is there any further relevant information you would like to provide? |

** To be updated, is discharged or at 30 days after sample date, whichever is sooner
